# Risk of Adverse Events Following Monovalent Third or Booster Dose of COVID-19 mRNA Vaccination in U.S. Adults Ages 18 Years and Older

**DOI:** 10.1101/2024.02.20.24303089

**Authors:** Azadeh Shoaibi, Kathryn Matuska, Patricia C. Lloyd, Hui Lee Wong, Joann F. Gruber, Tainya C. Clarke, Sylvia Cho, Emily Lassman, Hai Lyu, Rowan McEvoy, Zhiruo Wan, Mao Hu, Sandia Akhtar, Yixin Jiao, Yoganand Chillarige, Daniel Beachler, Alex Secora, Nandini Selvam, Djeneba Audrey Djibo, Cheryl N McMahill Walraven, John D. Seeger, Kandace L. Amend, Jennifer Song, Robin Clifford, Jeffrey A. Kelman, Richard A. Forshee, Steven A. Anderson

**Affiliations:** US Food and Drug Administration, Silver Spring, MD, USA; Acumen LLC, Burlingame, CA, USA; Carelon Research, Wilmington, DE, USA; IQVIA, Falls Church, VA, USA; CVS Health/Aetna, Blue Bell, Pennsylvania, USA; Optum Epidemiology, Boston, MA, USA; Centers for Medicare & Medicaid Services, Washington, DC, USA

**Keywords:** COVID-19 mRNA vaccines, monovalent booster, Pfizer-BioNTech, Moderna, Medicare, COVID-19 vaccine safety

## Abstract

**Background:** The U.S. FDA authorized the monovalent third primary series or booster doses of COVID-19 mRNA vaccines in August 2021 for persons 18 years and older. Monitoring of outcomes following updated authorizations is critical to evaluate vaccine safety and can provide early detection of rare adverse events (AEs) not identified in pre-licensure trials.

**Methods:** We evaluated the risk of 17 AEs following third doses of COVID-19 mRNA vaccines from August 2021 through early 2022 among adults aged 18-64 years in three commercial databases (Optum, Carelon Research, CVS Health) and adults aged >65 years in Medicare Fee-For-Service. We compared observed AE incidence rates to historical (expected) rates prior to the pandemic, estimated incidence rate ratios (IRRs) for the Medicare database and pooled IRR across the three commercial databases. Analyses were also stratified by prior history of COVID-19 diagnosis. Estimates exceeding a pre-defined threshold were considered statistical signals.

**Results:** Four AEs met the threshold for statistical signals for BNT162b2 and mRNA-1273 vaccines including Bell’s Palsy and pulmonary embolism in Medicare, and anaphylaxis and myocarditis/pericarditis in commercial databases. Nine AEs and three AEs signaled among adults with and without prior COVID-19 diagnosis, respectively.

**Conclusions:** This early monitoring study identified statistical signals for AEs following third doses of COVID-19 mRNA vaccination. Since this method is intended for screening purposes and generates crude results, results do not establish a causal association between the vaccines and AEs. FDA’s public health assessment remains consistent that the benefits of COVID-19 vaccination outweigh the risks of vaccination.

## Introduction

In the United States (U.S.), the BNT162b2 (Pfizer-BioNTech) and mRNA-1273 (Moderna) mRNA vaccines are available to prevent severe Coronavirus disease 2019 (COVID-19).^1,2^ For monovalent formulations, third doses as primary series doses were recommended for persons with immunocompromising conditions as of August 12, 2021, and a first booster dose (outside of the primary series) was authorized through Emergency Use Authorization (EUA) by the U.S. Food and Drug Administration (FDA) for everyone aged 18 years and older from November 19, 2021 to April 18, 2023. Statistical signals have been observed in active surveillance of the monovalent mRNA vaccines.^3–5^ Thus, the FDA Biologics Effectiveness and Safety (BEST) Initiative, an active surveillance program for post-market surveillance of biologic products, conducted this study to investigate the safety of COVID-19 mRNA vaccines among individuals aged 18 to 64 years in the commercially insured population and individuals aged 65 years and older in the Medicare population by evaluating the rates of 17 adverse events (AEs) of interest following third doses by comparing them to historical control (expected) rates for their respective AEs. Results from this signal detection study do not establish a causal association between the vaccines and AEs.

## Methods

### Data Sources

This study used administrative claims data from the Centers for Medicare & Medicaid Services (CMS) Medicare (Fee-For-Service) database and from three commercial health insurance databases: CVS Health, Optum pre-adjudicated claims, and Carelon Research (Supplemental Table 1). The commercial data sources were supplemented with Immunization Information System (IIS) data from local and state jurisdictions to increase the capture of vaccinations.^6^

### Study Population and Period

The study population included a commercially insured population (aged 18-64 years) and the publicly insured Medicare Fee-For-Service population (aged 65 years and older) who received a COVID-19 mRNA third dose vaccination from the study start date of August 12, 2021, when the FDA authorized an additional vaccine dose for certain immunocompromised individuals,^7^ through database-specific study end dates. Data are included through April 2022 for Optum and Carelon, February 2022 for CVS Health, and March 2022 for Medicare enrollees. For inclusion in AE-specific analyses, persons must have been continuously enrolled in a medical health insurance plan from the start of the AE-specific clean window through the date of their third COVID-19 vaccine dose. This AE-specific pre-vaccination clean window was defined as a specific number of days before vaccination date when a patient must not have had the AE (Table 1).

**Table 1.**
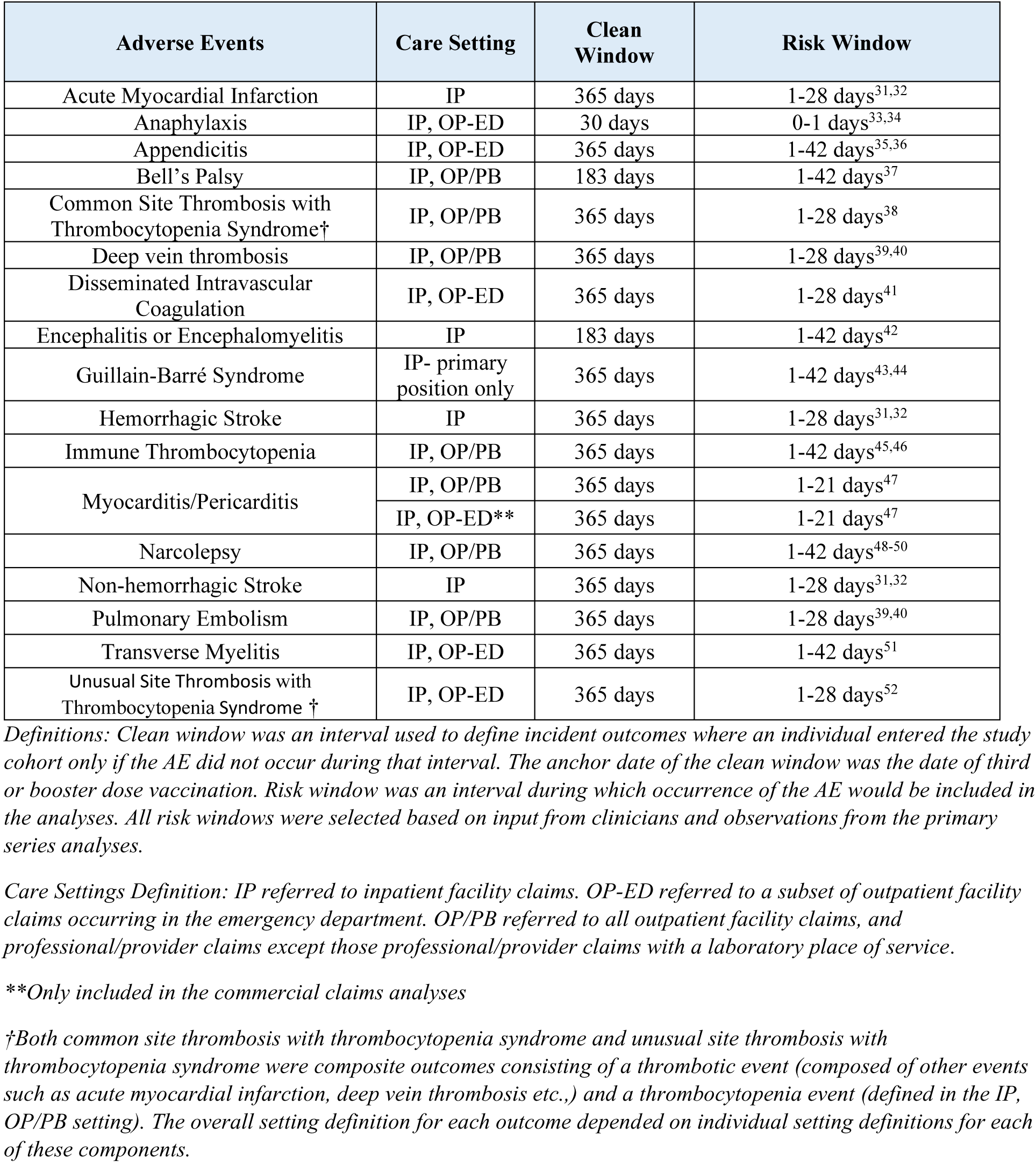
Study Specifications: Adverse Events (AEs), Care Settings, Clean Windows, and Risk Windows for the Medicare and Commercial Databases.

| Adverse Events | Care Setting | Clean Window | Risk Window |
| --- | --- | --- | --- |
| Acute Myocardial Infarction | IP | 365 days | 1-28 days<sup>31,32</sup> |
| Anaphylaxis | IP, OP-ED | 30 days | 0-1 days<sup>33,34</sup> |
| Appendicitis | IP, OP-ED | 365 days | 1-42 days<sup>35,36</sup> |
| Bell's Palsy | IP, OP/PB | 183 days | 1-42 days<sup>37</sup> |
| Common Site Thrombosis with Thrombocytopenia Syndrome† | IP, OP/PB | 365 days | 1-28 days<sup>38</sup> |
| Deep vein thrombosis | IP, OP/PB | 365 days | 1-28 days<sup>39,40</sup> |
| Disseminated Intravascular Coagulation | IP, OP-ED | 365 days | 1-28 days<sup>41</sup> |
| Encephalitis or Encephalomyelitis | IP | 183 days | 1-42 days<sup>42</sup> |
| Guillain-Barré Syndrome | IP- primary position only | 365 days | 1-42 days<sup>43,44</sup> |
| Hemorrhagic Stroke | IP | 365 days | 1-28 days<sup>31,32</sup> |
| Immune Thrombocytopenia | IP, OP/PB | 365 days | 1-42 days<sup>45,46</sup> |
| Myocarditis/Pericarditis | IP, OP/PB | 365 days | 1-21 days<sup>47</sup> |
|  | IP, OP-ED** | 365 days | 1-21 days<sup>47</sup> |
| Narcolepsy | IP, OP/PB | 365 days | 1-42 days<sup>48-50</sup> |
| Non-hemorrhagic Stroke | IP | 365 days | 1-28 days<sup>31,32</sup> |
| Pulmonary Embolism | IP, OP/PB | 365 days | 1-28 days<sup>39,40</sup> |
| Transverse Myelitis | IP, OP-ED | 365 days | 1-42 days<sup>51</sup> |
| Unusual Site Thrombosis with Thrombocytopenia Syndrome † | IP, OP-ED | 365 days | 1-28 days<sup>52</sup> |

*Definitions: Clean window was an interval used to define incident outcomes where an individual entered the study cohort only if the AE did not occur during that interval. The anchor date of the clean window was the date of third or booster dose vaccination. Risk window was an interval during which occurrence of the AE would be included in the analyses. All risk windows were selected based on input from clinicians and observations from the primary series analyses.*
*Care Settings Definition: IP referred to inpatient facility claims. OP-ED referred to a subset of outpatient facility claims occurring in the emergency department. OP/PB referred to all outpatient facility claims, and professional/provider claims except those professional/provider claims with a laboratory place of service.*
*\*\*Only included in the commercial claims analyses*
*†Both common site thrombosis with thrombocytopenia syndrome and unusual site thrombosis with thrombocytopenia syndrome were composite outcomes consisting of a thrombotic event (composed of other events such as acute myocardial infarction, deep vein thrombosis etc.,) and a thrombocytopenia event (defined in the IP, OP/PB setting). The overall setting definition for each outcome depended on individual setting definitions for each of these components.*

### Exposure and Follow-Up

Exposure was defined as the receipt of a third dose of the BNT162b2 COVID-19 or the mRNA-1273 COVID-19 vaccine (Supplemental Table 2). Vaccinations were identified using brand and dose-specific Current Procedural Terminology (CPT)/Healthcare Common Procedure Coding System (HCPCS) codes or National Drug Codes (NDCs) in the professional, outpatient institutional, inpatient, or prescription drug care settings,^8^ and were identified by CVX (vaccine administered) product codes in IIS data sources.

Dose assignment was based on the chronological order in which vaccinations were observed for each person. Third doses were restricted to doses following a two-dose primary series of mRNA vaccines. If a patient was administered more than 3 doses of BNT162b2 or mRNA-1273 vaccine, follow-up time was censored at the time of the fourth dose administration.

Patients who received heterologous vaccine brands for the primary series and third dose and persons who received the first booster dose of Ad26.COV2.S (Janssen) vaccine were included in the descriptive analyses but not in the inferential analyses due to small sample size. First booster doses of the Ad26.COV2.S vaccine were restricted to doses following a one-dose primary series, and follow-up time was censored at the time of administration if a patient was administered more than 2 doses of Ad26.COV2.S.

Follow-up time began on the day after vaccination and was censored at death, disenrollment, end of AE-specific risk window (Table 1), end of study period, a subsequent COVID-19 vaccine dose, or at AE occurrence, whichever came first.

### Adverse Events of Interest

Claims-based AE algorithms, pre-vaccination clean windows, and post-vaccination risk windows were selected based on literature review and clinical consultation. All AEs (Table 1) were identified using International Classification of Diseases, Tenth Revision, Clinical Modification (ICD-10-CM) diagnosis codes.^8^ Claims from inpatient facilities (IP), emergency department encounters in outpatient facilities (OP-ED), and all outpatient facilities and individual providers or professionals (OP/PB) were used to identify AEs.

### Statistical Analyses

The vaccine dose count was summarized by brand for each AE (Table 2). The count and proportion of heterologous vaccine brand administration of third doses was also summarized.

**Table 2.**
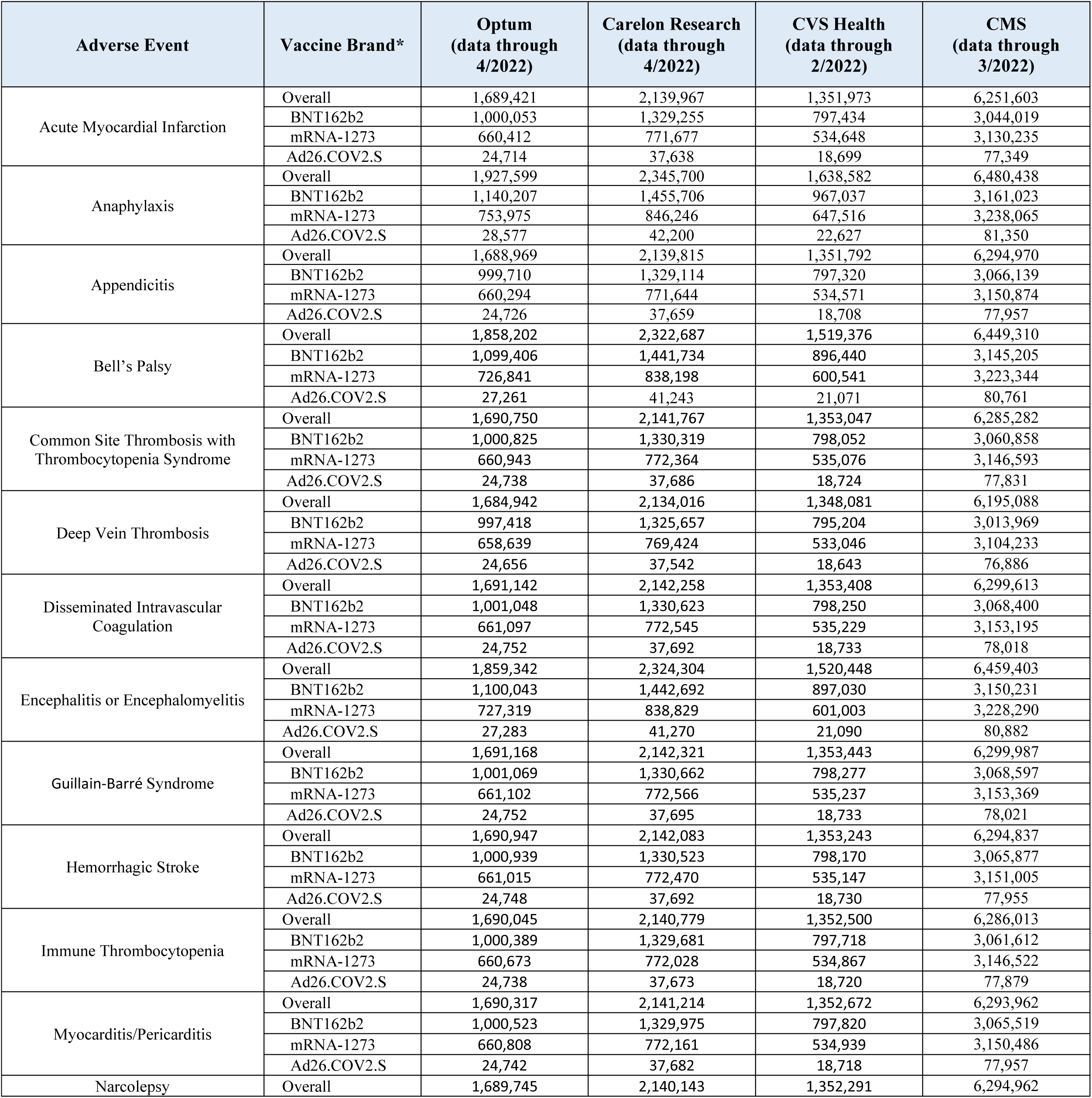
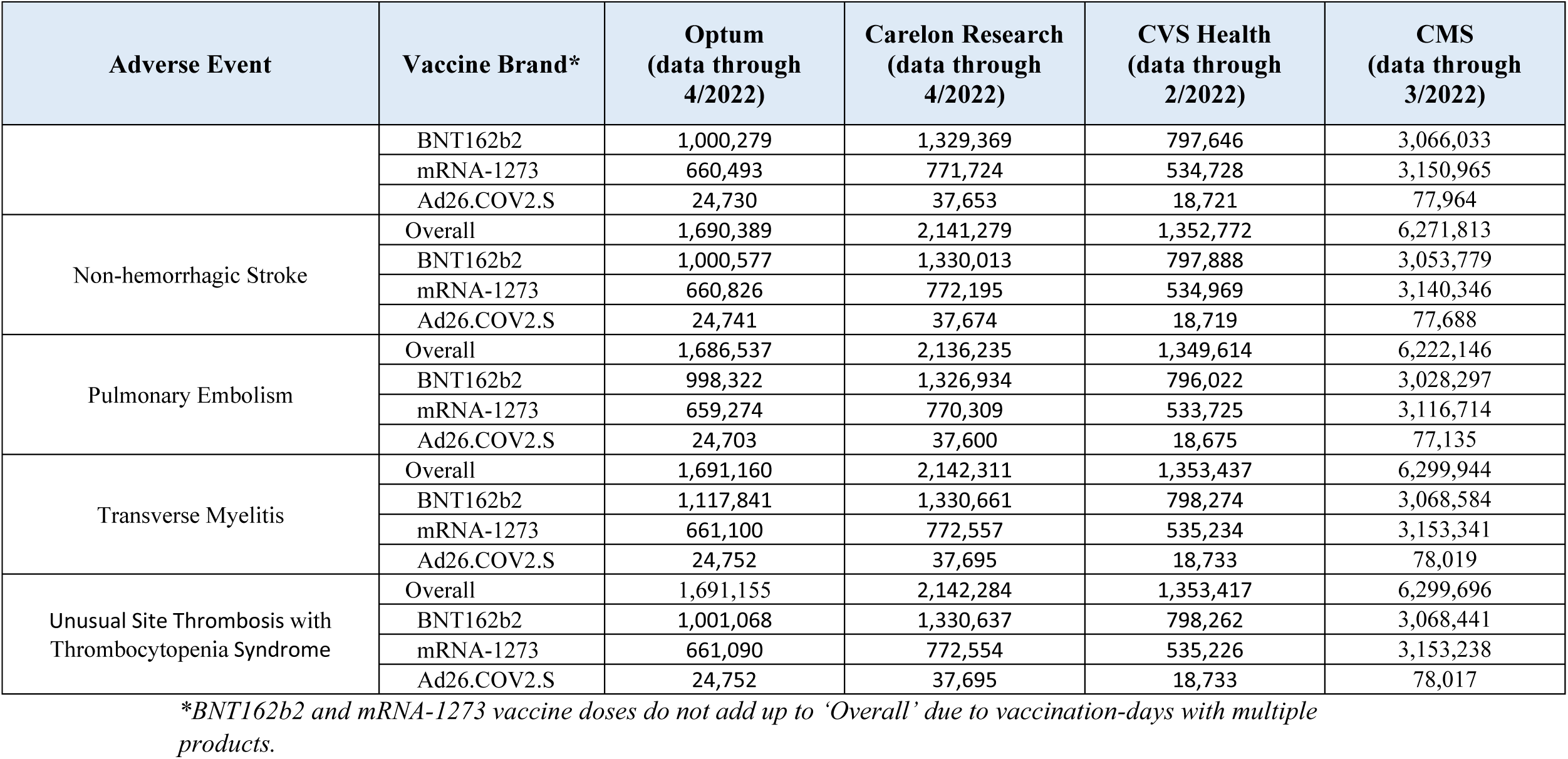
Number of Third COVID-19 Vaccine Doses Administered Stratified by Adverse Event and Vaccine Brand.

This study was designed as a retrospective cohort analysis with a historical comparator group. The primary analysis compared the observed rate of the AE of interest in the post-vaccination risk window to the rate in a database-specific historical control population, and incidence rate ratios (IRR) were estimated. The observed rate of each AE was adjusted based on observation delay for that outcome in 2019 using a similar approach as described in the literature for partially accrued data.^9^ We calculated database-specific comparator rates as the annual AE rates within the overall and influenza-vaccinated populations in the pre-COVID-19 (2017-2019) and during-COVID-19 pandemic (June 2020-December 10, 2020) periods. The comparator rate used for the analysis was selected to enhance the sensitivity of the test for signal detection. If annual rates in the historical period differed, as assessed by comparing the 95% confidence intervals (CI), we selected the lowest rate; otherwise, the median annual rate was selected. Pre-COVID-19 rates from 2019 were selected for most outcomes; however, when the during-COVID-19 rate (June-December 10, 2020) did not return to pre-COVID-19 levels, rates from 2020 were selected.^8^ There were no adjustments for baseline clinical characteristics. Comparator rates were generated stratified by age and sex when sufficient counts in the respective strata were available in all databases. Also, comparator rates were generated by nursing home residency status and race in the Medicare population only.

Analyses for myocarditis/pericarditis were stratified by care setting (all settings and IP/OP-ED settings) and age (18-45 years and 46-64 years) in the commercially insured population to more granularly assess risk based on prior evidence regarding increased risk of myocarditis/pericarditis following COVID-19 mRNA vaccination particularly amongst younger males.^10–22^

For each potential AE, Poisson models were used to estimate the incidence rate (IR) per 100,000 person-years with 95% CI and the IRR of observed IR vs. expected IR with 95% CI. One-sided tests were conducted with an overall alpha of 2.5% and with testing margins selected for each AE based on clinical significance and expert guidance to avoid detection of false-positive results and characterization of non-clinically meaningful, minimal risk increases. In the commercially insured population, the null hypotheses were IRR = 1.5 for anaphylaxis, transverse myelitis, and myocarditis/pericarditis; 2.5 for encephalitis or encephalomyelitis, narcolepsy, and Guillain-Barré syndrome (GBS); and 1.25 for all other outcomes. To detect any increase in risk in the Medicare population, in which outcomes are more common, more sensitive tests were conducted where the null hypothesis was IRR = 1 for all AEs. A statistical signal occurred if the observed IRR exceeded the null hypothesis and the null hypothesis was not within the IRR 95% CI which was used as a threshold for determining whether the result was likely to occur due to chance. Statistical testing was only conducted for individuals who received the same vaccine brand for primary series and third doses.

To account for prior COVID-19 infection as a potential confounder, we conducted a secondary analysis for each AE stratified by history of COVID-19 diagnosis identified by diagnosis of COVID-19 in any care setting (identified by U07.1 ICD-10-CM code) on or following April 1, 2020.

IRs and IRRs were generated for the Medicare database separately and pooled across the three commercial databases using meta-analysis with a random-effects and fixed-effects models. Between-study heterogeneity was assessed using a *I*^2^ statistic.^23^

## Results

### Descriptive Analyses

Across the commercial insurance databases, 3,803,442 beneficiaries received BNT162b2 and 2,400,502 beneficiaries received mRNA-1273 third dose vaccines. The count of vaccine doses was summarized by brand for each AE for all COVID-19 vaccines (Table 2) and AE-specific analyses (Supplemental Table 3). A large proportion of third doses was mRNA vaccines; we did not conduct inferential analyses on the Ad26.COV2.S vaccine due to the small number of doses administered. Approximately 64% - 83% of vaccinations across brands in commercial databases were eligible for the analysis and contributed to an AE-specific cohort. In the Medicare database, 3,336,862 beneficiaries received BNT162b2 and 3,399,048 beneficiaries received mRNA-1273 third dose vaccines; more than 90% of these vaccinations across brands were eligible for the analysis and contributed to an AE-specific cohort (Supplemental Table 3). Less than 13% of cases with an observed AE across databases had an associated prior COVID-19 diagnosis (Supplemental Table 4, Supplemental Table 5). Most people received the same vaccine brand for primary series and third doses (Supplemental Table 6).

### Results of Primary and Secondary Analyses – Persons 18 to 64 years

Among the 17 AEs of interest evaluated in the commercially insured population, two AEs met the threshold for a statistical signal in primary analyses. A signal was observed post-BNT162b2 third dose vaccination for anaphylaxis (IRR = 4.92, 95% CI = 2.20–10.98). Following mRNA-1273 third dose vaccination, signals were observed for anaphylaxis (IRR = 5.70, 95% CI = 1.69– 19.21) and myocarditis/pericarditis: IP/OP-ED (18-45 years) (IRR = 3.38, 95% CI = 1.51–7.58) (Table 3).

**Table 3.** Incidence Rate Ratio (IRR) and 95% Confidence Interval (CI) of Adverse Events Following a Third Dose of COVID-19 mRNA Vaccine Among Adults 18–64 Years Old Enrolled in Commercial Insurance (Meta-Analyzed across Commercial Databases)

| Adverse Events | Vaccine Brand | No. Observed Cases | IRR (95% CI) |
| --- | --- | --- | --- |
| Acute Myocardial Infarction | BNT162b2 | 186 | 0.82 (0.71 – 0.95) |
|  | mRNA-1273 | 140 | 0.97 (0.82 – 1.14) |
| Anaphylaxis | BNT162b2 | <11 | **4.92\* (2.20 – 10.98)** |
|  | mRNA-1273 | <11 | **5.70\* (1.69 – 19.21)** |
| Appendicitis | BNT162b2 | 295 | 0.95 (0.73 – 1.25) |
|  | mRNA-1273 | 164 | 0.89 (0.66 – 1.21) |
| Bell’s Palsy | BNT162b2 | 331 | 0.97 (0.83 – 1.13) |
|  | mRNA-1273 | 173 | 0.84 (0.62 – 1.13) |
| Common Site Thrombosis with Thrombocytopenia Syndrome | BNT162b2 | 43 | 0.82 (0.61 – 1.11) |
|  | mRNA-1273 | 25 | 0.68 (0.23 – 2.04) |
| Deep Vein Thrombosis | BNT162b2 | 357 | 0.76 (0.66 – 0.88) |
|  | mRNA-1273 | 258 | 0.87 (0.77 – 0.99) |
| Disseminated Intravascular Coagulation | BNT162b2 | <11 | 0.94 (0.49 – 1.80) |
|  | mRNA-1273 | <11 | 1.22 (0.54 – 2.78) |
| Encephalitis or Encephalomyelitis | BNT162b2 | 12 | 2.28 (0.96 – 5.43) |
|  | mRNA-1273 | <11 | 1.28 (0.53 – 3.09) |
| Guillain-Barré Syndrome | BNT162b2 | <11 | 0.82 (0.37 – 1.84) |
|  | mRNA-1273 | <11 | 2.75 (0.76 – 9.95) |
| Hemorrhagic Stroke | BNT162b2 | 32 | 0.92 (0.48 – 1.74) |
|  | mRNA-1273 | 17 | 0.85 (0.53 – 1.34) |
| Immune Thrombocytopenia | BNT162b2 | 91 | 1.03 (0.70 – 1.52) |
|  | mRNA-1273 | 51 | 0.99 (0.65 – 1.50) |
| Myocarditis/Pericarditis: All Care Settings (18-45 years of age) | BNT162b2 | 41 | 1.95 (1.38 – 2.75) |
|  | mRNA-1273 | 24 | 2.19 (1.08 – 4.48) |
| Myocarditis/Pericarditis: All Care Settings (46-64 years of age) | BNT162b2 | 38 | 1.23 (0.90 – 1.68) |
|  | mRNA-1273 | 34 | 1.69 (1.21 – 2.35) |
| Myocarditis/Pericarditis: IP/OP-ED (18-45 years of age) | BNT162b2 | 15 | 2.31 (1.19 – 4.51) |
|  | mRNA-1273 | 11 | **3.38\* (1.51 – 7.58)** |
| Myocarditis/Pericarditis: IP/OP-ED (46-64 years of age) | BNT162b2 | <11 | 1.01 (0.52 – 1.93) |
|  | mRNA-1273 | 12 | 2.38 (1.24 – 4.56) |
| Narcolepsy | BNT162b2 | 96 | 0.82 (0.59 – 1.13) |
|  | mRNA-1273 | 57 | 0.89 (0.62 – 1.27) |
| Non-hemorrhagic Stroke | BNT162b2 | 97 | 0.8 (0.55 – 1.17) |
|  | mRNA-1273 | 65 | 0.82 (0.64 – 1.04) |
| Pulmonary Embolism | BNT162b2 | 309 | 1.01 (0.91 – 1.13) |
|  | mRNA-1273 | 181 | 0.97 (0.81 – 1.15) |
| Transverse Myelitis | BNT162b2 | <11 | 0.98 (0.29 – 3.31) |
|  | mRNA-1273 | <11 | 1.23 (0.36 – 4.14) |
| Unusual Site Thrombosis with Thrombocytopenia Syndrome | BNT162b2 | <11 | 0.66 (0.24 – 1.78) |
|  | mRNA-1273 | <11 | 0.94 (0.28 – 3.16) |

*\*Indicates the adverse event for the specific vaccine brand has signaled in the analyses.*
*Cell sizes 1-10 were masked for confidentiality.*
*Care Settings Definition: IP referred to inpatient facility claims. OP-ED referred to a subset of outpatient facility claims occurring in the emergency department. OP/PB referred to all outpatient facility claims, and professional/provider claims except those professional/provider claims with a laboratory place of service.*

In the secondary analyses stratified by prior COVID-19 diagnosis, statistical signals were observed in the population with a prior COVID-19 diagnosis following BNT162b2 third dose vaccination for anaphylaxis (IRR= 37.26, 95% CI = 6.69–207.58); encephalitis or encephalomyelitis (IRR= 26.85, 95% CI = 7.43 – 96.99); and unusual site TTS (IRR= 8.43, 95% CI = 1.51–46.96) (Supplemental Table 4). The following statistical signals were observed following mRNA-1273 third dose vaccination: myocarditis/pericarditis, all settings for 18-45 years (IRR= 7.22, 95% CI = 2.58–20.19) and 46-64 years (IRR= 6.61, 95% CI = 3.26–13.41); myocarditis/pericarditis, IP/OP-ED settings for 46-64 years (IRR= 11.35, 95% CI = 4.06–31.74); common site thrombosis with thrombocytopenia (TTS) (IRR= 5.83, 95% CI = 2.28–14.92); and GBS (IRR= 18.19, 95% CI = 3.26–101.35).

In the population without a prior history of COVID-19 diagnosis, a signal was observed post-BNT162b2 third dose vaccination for anaphylaxis (IRR= 6.21, 95% CI = 2.50–15.39). In addition, signals were observed post-mRNA-1273 third dose vaccinations for myocarditis/pericarditis, IP/OP-ED settings for 18-45 years (IRR= 3.63, 95% CI = 1.62–8.15); and anaphylaxis (IRR= 6.15, 95% CI = 1.82–20.71) (Supplemental Table 4).

### Results of Primary and Secondary Analyses –Persons 65 years and older

Statistical signals were observed for two AEs in the primary analysis in the Medicare population following third dose vaccinations. Statistical signals were identified following BNT162b2 third dose vaccinations for Bell’s Palsy (BP) (IRR = 1.11, 95% CI = 1.03 – 1.19) and pulmonary embolism (PE) (IRR = 1.05, 95% CI = 1.001 – 1.100) (Table 4). No signals were identified following mRNA-1273 third dose vaccinations (Table 4).

**Table 4.**
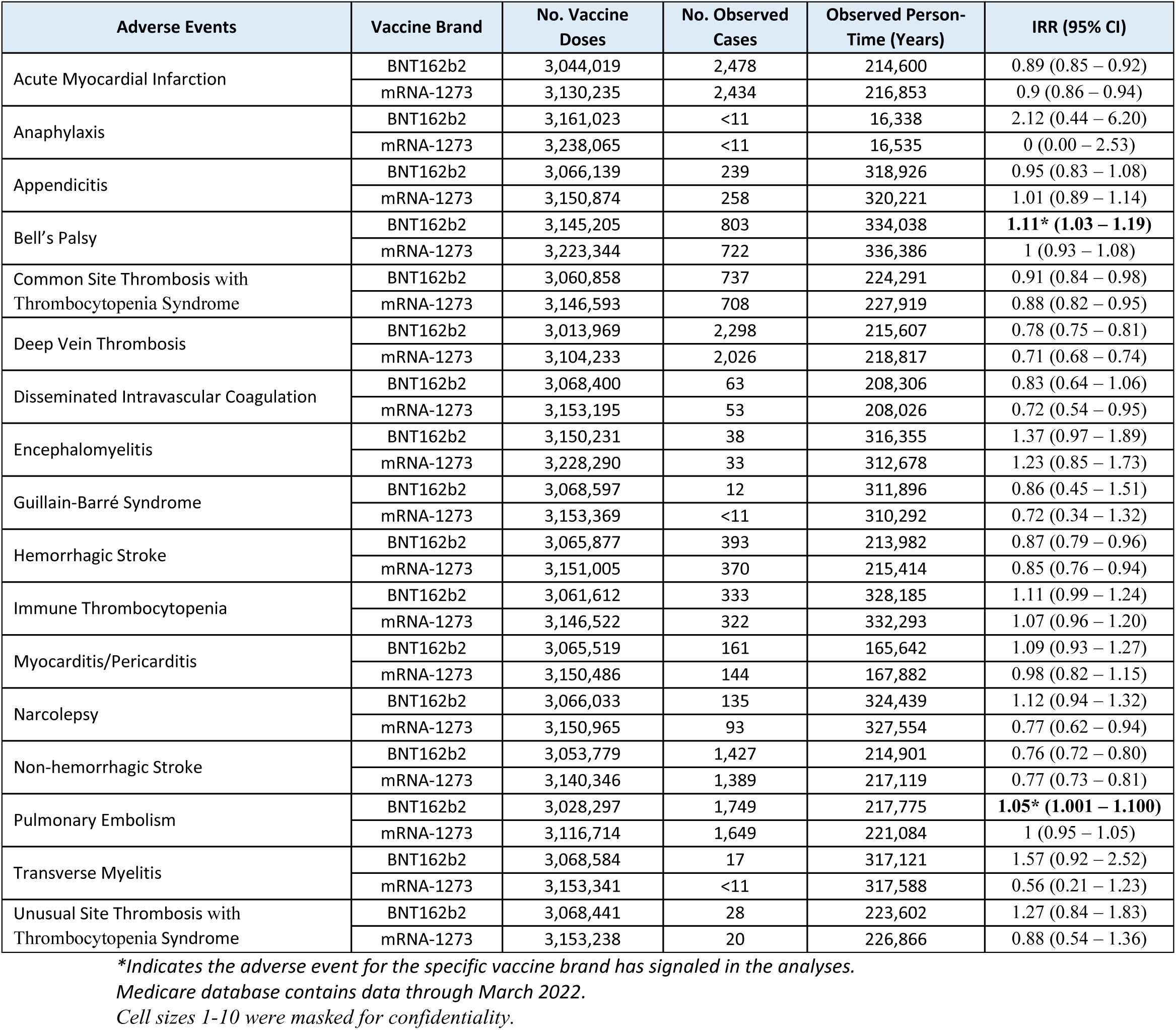
Incidence Rate Ratio (IRR) and 95% Confidence Interval (CI) of Adverse Events Following a Third Dose of COVID-19 mRNA Vaccine Among Adults 65 Years and Older Enrolled in Medicare.

| Adverse Events | Vaccine Brand | No. Vaccine Doses | No. Observed Cases | Observed Person-Time (Years) | IRR (95% CI) |
| --- | --- | --- | --- | --- | --- |
| Acute Myocardial Infarction | BNT162b2 | 3,044,019 | 2,478 | 214,600 | 0.89 (0.85 – 0.92) |
|  | mRNA-1273 | 3,130,235 | 2,434 | 216,853 | 0.9 (0.86 – 0.94) |
| Anaphylaxis | BNT162b2 | 3,161,023 | <11 | 16,338 | 2.12 (0.44 – 6.20) |
|  | mRNA-1273 | 3,238,065 | <11 | 16,535 | 0 (0.00 – 2.53) |
| Appendicitis | BNT162b2 | 3,066,139 | 239 | 318,926 | 0.95 (0.83 – 1.08) |
|  | mRNA-1273 | 3,150,874 | 258 | 320,221 | 1.01 (0.89 – 1.14) |
| Bell's Palsy | BNT162b2 | 3,145,205 | 803 | 334,038 | **1.11\* (1.03 – 1.19)** |
|  | mRNA-1273 | 3,223,344 | 722 | 336,386 | 1 (0.93 – 1.08) |
| Common Site Thrombosis with Thrombocytopenia Syndrome | BNT162b2 | 3,060,858 | 737 | 224,291 | 0.91 (0.84 – 0.98) |
|  | mRNA-1273 | 3,146,593 | 708 | 227,919 | 0.88 (0.82 – 0.95) |
| Deep Vein Thrombosis | BNT162b2 | 3,013,969 | 2,298 | 215,607 | 0.78 (0.75 – 0.81) |
|  | mRNA-1273 | 3,104,233 | 2,026 | 218,817 | 0.71 (0.68 – 0.74) |
| Disseminated Intravascular Coagulation | BNT162b2 | 3,068,400 | 63 | 208,306 | 0.83 (0.64 – 1.06) |
|  | mRNA-1273 | 3,153,195 | 53 | 208,026 | 0.72 (0.54 – 0.95) |
| Encephalomyelitis | BNT162b2 | 3,150,231 | 38 | 316,355 | 1.37 (0.97 – 1.89) |
|  | mRNA-1273 | 3,228,290 | 33 | 312,678 | 1.23 (0.85 – 1.73) |
| Guillain-Barré Syndrome | BNT162b2 | 3,068,597 | 12 | 311,896 | 0.86 (0.45 – 1.51) |
|  | mRNA-1273 | 3,153,369 | <11 | 310,292 | 0.72 (0.34 – 1.32) |
| Hemorrhagic Stroke | BNT162b2 | 3,065,877 | 393 | 213,982 | 0.87 (0.79 – 0.96) |
|  | mRNA-1273 | 3,151,005 | 370 | 215,414 | 0.85 (0.76 – 0.94) |
| Immune Thrombocytopenia | BNT162b2 | 3,061,612 | 333 | 328,185 | 1.11 (0.99 – 1.24) |
|  | mRNA-1273 | 3,146,522 | 322 | 332,293 | 1.07 (0.96 – 1.20) |
| Myocarditis/Pericarditis | BNT162b2 | 3,065,519 | 161 | 165,642 | 1.09 (0.93 – 1.27) |
|  | mRNA-1273 | 3,150,486 | 144 | 167,882 | 0.98 (0.82 – 1.15) |
| Narcolepsy | BNT162b2 | 3,066,033 | 135 | 324,439 | 1.12 (0.94 – 1.32) |
|  | mRNA-1273 | 3,150,965 | 93 | 327,554 | 0.77 (0.62 – 0.94) |
| Non-hemorrhagic Stroke | BNT162b2 | 3,053,779 | 1,427 | 214,901 | 0.76 (0.72 – 0.80) |
|  | mRNA-1273 | 3,140,346 | 1,389 | 217,119 | 0.77 (0.73 – 0.81) |
| Pulmonary Embolism | BNT162b2 | 3,028,297 | 1,749 | 217,775 | **1.05\* (1.001 – 1.100)** |
|  | mRNA-1273 | 3,116,714 | 1,649 | 221,084 | 1 (0.95 – 1.05) |
| Transverse Myelitis | BNT162b2 | 3,068,584 | 17 | 317,121 | 1.57 (0.92 – 2.52) |
|  | mRNA-1273 | 3,153,341 | <11 | 317,588 | 0.56 (0.21 – 1.23) |
| Unusual Site Thrombosis with Thrombocytopenia Syndrome | BNT162b2 | 3,068,441 | 28 | 223,602 | 1.27 (0.84 – 1.83) |
|  | mRNA-1273 | 3,153,238 | 20 | 226,866 | 0.88 (0.54 – 1.36) |

*\*Indicates the adverse event for the specific vaccine brand has signaled in the analyses.*
*Medicare database contains data through March 2022.*
*Cell sizes 1-10 were masked for confidentiality.*

In the secondary analyses stratified by prior COVID-19 diagnosis, five AEs met the threshold for a statistical signal. In the population with a prior COVID-19 diagnosis, signals were observed for PE following BNT162b2 (IRR = 1.36, 95% CI = 1.17–1.58) and mRNA-1273 third dose vaccinations (IRR = 1.43, 95% CI = 1.20–1.69); acute myocardial infarction (AMI) following BNT162b2 (IRR = 1.15, 95% CI = 1.02–1.29) and mRNA-1273 third dose vaccinations (IRR = 1.33, 95% CI = 1.16–1.52). In addition, following BNT162b2 third dose vaccination, signals were observed for immune thrombocytopenia (ITP) (IRR = 1.66, 95% CI = 1.17–2.29) and myocarditis/pericarditis (IRR = 1.89, 95% CI = 1.18–2.86).

In the population without a prior history of COVID-19 diagnosis, a signal was observed for BP (IRR = 1.11, 95% CI = 1.03–1.19) following BNT162b2 third dose vaccination (Supplemental Table 6). While more AEs of interest with elevated risk were observed in the secondary analysis compared to the primary analysis, smaller sample sizes reflected wider 95% CI estimates (Supplemental Tables 4–5).

## Discussion

In this large population-based study we investigated the safety of third doses of COVID-19 mRNA vaccines by examining the risk of 17 AEs following vaccination in a signal detection study as the first step in the safety surveillance process.

The primary analyses of the study found statistical signals for four AEs. Myocarditis/pericarditis and anaphylaxis have been reported to be associated with the primary doses of vaccines.^3,4,8,10–22,24^ There is a high level of outcome misclassification (a non-case being identified as a case or vice versa) associated with the detection of BP cases and hence uncertainty with its signal as results from a medical record review (MRR) of a sample of BP cases in our other COVID-19 vaccine studies showed a low positive predictive value (PPV) (<20%, data not shown) of the claims-based diagnosis codes used to identify cases indicating that most cases resulting in the signal were not true cases. The PE signal was marginal and similar to inconsistent results detected in previous COVID-19 vaccine safety studies where a protective effect following third doses of both mRNA brands and a small statistically significant elevated risk following BNT162b2 primary series were observed for PE.^24^

Among those with a prior COVID-19 diagnosis, nine AEs signaled. Most of these AEs have been reported to be associated with COVID-19 infection itself.^25–27^ In adults aged 18-64 years, all signals in populations with and without prior COVID-19 diagnosis arose from a small number of cases. Similar to BP, our MRR of ITP cases in other COVID-19 vaccines safety studies suggested a low PPV (<10%, data not shown) of the claims-based diagnosis codes used to identify cases indicating that most cases identified for inclusion were not true cases. Moreover, given the small number of cases, wide 95% CIs, and IRRs that are unadjusted for confounders, these signals carry a high level of uncertainty and should be interpreted cautiously.

In the population without a prior COVID-19 diagnosis, three AEs signaled. The BP signal may be explained by potential outcome misclassification, and the anaphylaxis and myocarditis/pericarditis signals are consistent with the current literature.^3,10–15^

This study has several strengths. The study included a large U.S. adult population covered by commercial insurance or Fee-For-Service Medicare. Because of the large sample size, the study had higher power to detect a smaller increase in risk of rare AEs after vaccination that may not be captured in pre-authorization clinical trials. Linkage of the commercial administrative claims databases to the IIS databases captured more COVID-19 vaccines administered. The retrospective cohort design with the use of historical comparator group allowed a rapid safety evaluation of vaccines as the first step in the active surveillance process.

There are several limitations in this study. Aggregate historical rates rather than individuals were used as the comparator which increases the potential for confounding and bias. Thus, the study results are crude or unadjusted for confounders, and do not establish a causal relationship between the vaccines and AEs. The stratified results should also be interpreted in the context of uncertainty and with caution. Although this study design was unable to disentangle the effect of prior COVID-19 infection and COVID-19 vaccines on the rates of AEs, several AEs which signaled in the population with a prior history of COVID-19 diagnosis are known to be associated with COVID-19 infection.^25–27^ Therefore, it is likely that a number of detected signals from these crude results may be associated with the prior history of COVID-19 diagnosis.

In this study, AEs were identified by diagnosis codes in claims databases which are subject to coding errors, and the study did not conduct MRR for all AEs to verify the true cases. Thus, outcome misclassification cannot be ruled out and could also bias results. The misclassification also applies to detection of prior COVID-19 infection in claims databases for the same reason as well as others such as use of home tests or not seeking medical care for the infection. Several identified signals arose from a very small number of AE cases. AE risk intervals were pre-specified based on literature review and clinical input, but their misspecification could lead to bias. The large sample size, inclusion of many AEs, and the number of statistical tests could increase the probability of false-positive results. Additionally, our study may not have captured all COVID-19 vaccines administered as some people may have received vaccines from sources that did not submit claims to insurance carriers, such as mass vaccination sites. Lastly, the results of this study may not be generalizable to the portion of the U.S. population who is uninsured or carries other types of health insurance.

Since the authorization or approval of the mRNA vaccines that were evaluated in this study, there have been additional FDA authorizations of COVID-19 vaccines. This study did not include second booster doses authorized for those 50 years and older or those 12 years and older with certain immunocompromised conditions (authorized March 29, 2022), third doses for children ages 5 to 11 years (authorized May 17, 2022), or bivalent booster doses for individuals 12 years and older (authorized August 31, 2022) as these vaccines were not yet authorized for use at the time this study was conducted.^1,28–30^ Active safety surveillance is ongoing to expand the monitoring of AEs to include individuals who receive these vaccinations.

In conclusion, this signal detection study detected multiple signals after exposure to the third dose of COVID-19 mRNA vaccines. The results of this study must be interpreted in the context of the study design, statistical methods, and their limitations; thus, some of the safety signals detected may be false positive results. While this study has major limitations, it contributes to the signal detection phase in the multi-step surveillance process for COVID-19 mRNA vaccines. FDA’s public health assessment remains consistent that the benefits of COVID-19 vaccination outweigh the risks of COVID-19 disease, and safety surveillance of new COVID-19 vaccines is ongoing.

## Tables

**Supplemental Table 1.** Description of Characteristics of Administrative Claims Data Sources Used in the Study.

| Data Source |  | Update frequency | Claims Data Lag | Enrollees |
| --- | --- | --- | --- | --- |
| Centers for Medicare & Medicaid Services (CMS) administrative claims data source | Medicare Fee-For-Service | Weekly | Approximately 80% data completeness in 1-3 months for inpatient, outpatient, and professional claims | ~48 million totals from 2017-2021<br>~33 million annually |
| Commercial claims data sources | CVS Health | Monthly | Approximately 80% data completeness in 3-4 months for inpatient claims, 2-3 months for outpatient claims, and 1-2 months for | ~37 million total from 2018-2021 ~22 million annually |
|  | Optum Pre-Adjudicated Claims | Bi-Weekly | Approximately 80% data completeness in 1-2 months for inpatient, outpatient, and professional claims | ~29 million total from 2017-2021 ~15-16 million annually |
|  | Carelon Research | Monthly | Approximately 80% data completeness in 2-3 months for inpatient claims and 1-2 months for outpatient and professional claims | ~46 million total from 2016-2021 ~24 million annually |

**Supplemental Table 2.** COVID-19 Vaccine Emergency Use Authorization (EUA) Dates and Dose Schedule Recommendations.

| Manufacturer | Name | Vaccine Authorization Dates<sup>1</sup> |  | Dosing Interval<sup>2,7,29,30,53</sup> |
| --- | --- | --- | --- | --- |
| Pfizer-BioNTech | BNT162b2<br>BNT162b2-BioNTech<br>COVID-19 Vaccine | 1<sup>st</sup> Dose | 12/11/2020 | - 21 days between doses 1, 2 |
|  |  | 2<sup>nd</sup> Dose | 12/11/2020 | - 28 days between doses 2, 3 |
|  |  | 3<sup>rd</sup> Dose | 8/12/2021 | - 5 months between primary series and booster dose |
|  |  | 1<sup>st</sup> Monovalent Booster | 9/22/2021 | - 4 months between first and second booster doses |
|  |  | 2<sup>nd</sup> Monovalent Booster | 3/29/2022 |  |
|  |  | Bivalent Booster Dose | 8/31/2022 |  |
| Moderna | mRNA-1273<br>COVID-19 Vaccine | 1<sup>st</sup> Dose | 12/18/2020 | - 28 days between doses 1, 2 |
|  |  | 2<sup>nd</sup> Dose | 12/18/2020 | - 28 days between doses 2, 3 |
|  |  | 3<sup>rd</sup> Dose | 8/12/2021 | - 5 months between primary series and booster dose |
|  |  | 1<sup>st</sup> Monovalent Booster | 10/20/2021 | - 4 months between first and second booster doses |
|  |  | 2<sup>nd</sup> Monovalent Booster | 3/29/2022 |  |
|  |  | Bivalent Booster Dose | 8/31/2022 |  |
| Janssen | Ad26.COV2.S<br>COVID-19 | 1<sup>st</sup> Dose | 2/27/2021 | - 2 months between primary series and booster dose |
|  |  | Monovalent Booster | 10/20/2022 |  |

**Supplemental Table 3.**
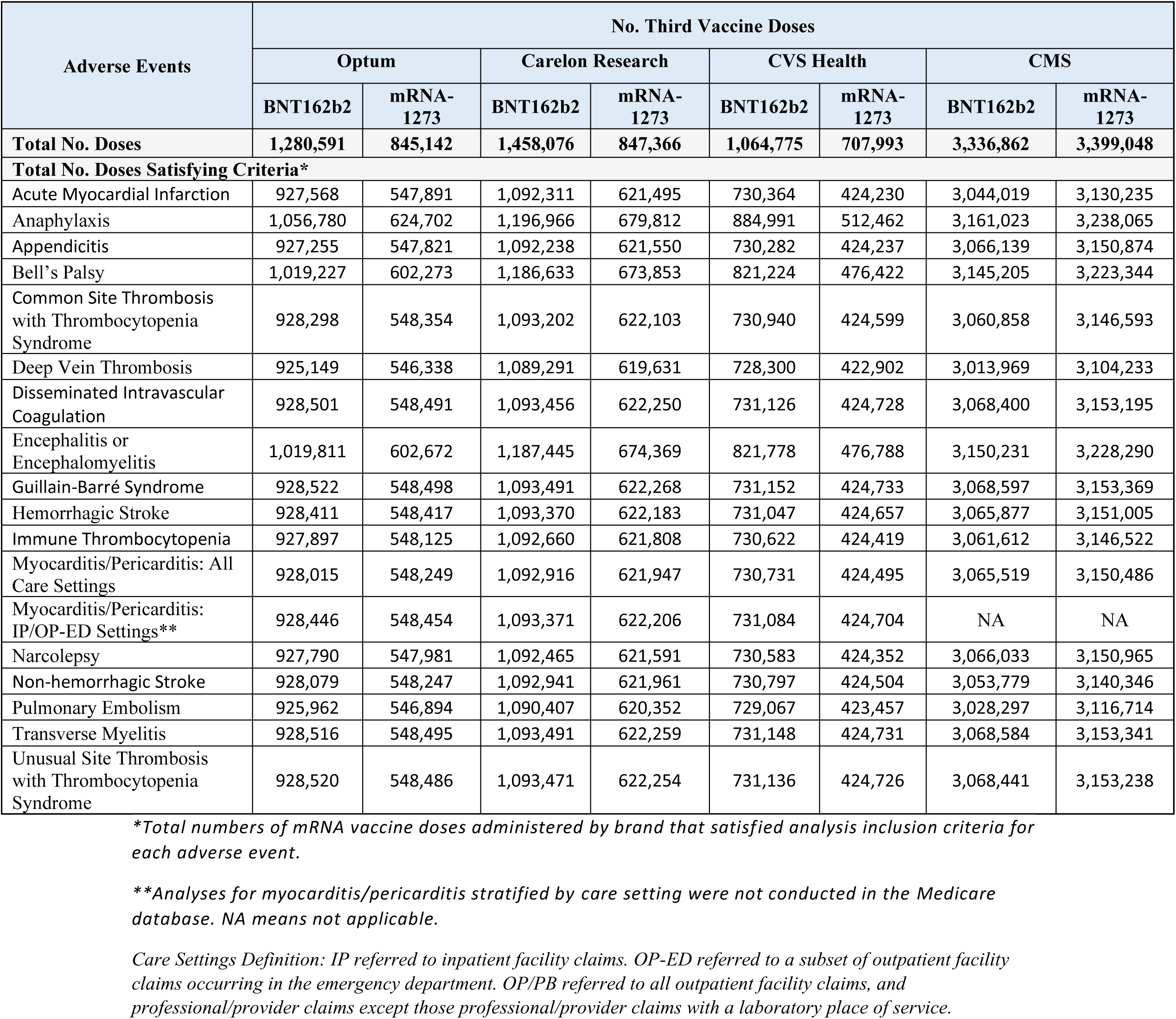
Number of Third COVID-19 mRNA Vaccine Doses Included in Adverse Event Specific Analyses.

**Supplemental Table 4.** Incidence Rate Ratio (IRR) and 95% Confidence Interval (CI) of Adverse Events Following a Third Dose of COVID-19 mRNA Vaccine Among Adults 18-64 Years Old Enrolled in Commercial Insurance Databases, Stratified by Prior COVID-19 Diagnosis, Meta-Analyzed for Commercial Claims Databases.

| Adverse Events | Vaccine Brand | With Prior COVID-19 Diagnosis |  | Without Prior COVID-19 Diagnosis |  |
| --- | --- | --- | --- | --- | --- |
|  |  | No. Observed Cases | IRR (95% CI) | No. Observed Cases | IRR (95% CI) |
| Acute Myocardial Infarction | BNT162b2 | 22 | 1.32 (0.88– 1.99) | 164 | 0.79 (0.68 – 0.92) |
|  | mRNA-1273 | 13 | 1.35 (0.70 – 2.57) | 127 | 0.95 (0.80 – 1.13) |
| Anaphylaxis | BNT162b2 | <11 | **37.26\* (6.69 – 207.58)** | <11 | **6.21\* (2.50 – 15.39)** |
|  | mRNA-1273 | 0 | NA | <11 | **6.15\* (1.82 – 20.71)** |
| Appendicitis | BNT162b2 | 29 | 1.22 (0.86 – 1.75) | 266 | 0.93 (0.71 – 1.23) |
|  | mRNA-1273 | 17 | 1.12 (0.28 – 4.46) | 147 | 0.87 (0.68 – 1.10) |
| Bell’s Palsy | BNT162b2 | 38 | 1.42 (0.83 – 2.45) | 293 | 0.93 (0.77 – 1.12) |
|  | mRNA-1273 | 17 | 0.95 (0.26 – 3.49) | 156 | 0.82 (0.64 – 1.04) |
| Common Site Thrombosis with Thrombocytopenia Syndrome | BNT162b2 | <11 | 1.01 (0.42 – 2.44) | 39 | 0.81 (0.60 – 1.11) |
|  | mRNA-1273 | <11 | **5.83\* (2.28 – 14.92)** | 21 | 0.66 (0.27 – 1.65) |
| Deep Vein Thrombosis | BNT162b2 | 46 | 1.27 (0.90 – 1.77) | 311 | 0.72 (0.63 – 0.81) |
|  | mRNA-1273 | 32 | 1.54 (0.96 – 2.47) | 226 | 0.83 (0.73 – 0.94) |
| Disseminated Intravascular Coagulation | BNT162b2 | <11 | 4.26 (0.76 – 23.74) | <11 | 0.95 (0.48 – 1.91) |
|  | mRNA-1273 | 0 | NA | <11 | 1.31 (0.58 – 2.99) |
| Encephalitis or Encephalomyelitis | BNT162b2 | <11 | **26.85\* (7.43 – 96.99)** | <11 | 1.98 (1.10 – 3.58) |
|  | mRNA-1273 | 0 | NA | <11 | 1.37 (0.57 – 3.31) |
| Guillain-Barré Syndrome | BNT162b2 | <11 | 4.91 (0.88 – 27.35) | <11 | 0.76 (0.32 – 1.84) |
|  | mRNA-1273 | <11 | **18.19\* (3.26 – 101.35)** | <11 | 1.49 (0.27 – 8.30) |
| Hemorrhagic Stroke | BNT162b2 | <11 | 1.7 (0.61 – 4.76) | 29 | 0.93 (0.54 – 1.62) |
|  | mRNA-1273 | <11 | 2.22 (0.66 – 7.48) | 15 | 0.8 (0.49 – 1.30) |
| Immune Thrombocytopenia | BNT162b2 | <11 | 1.14 (0.56 – 2.31) | 84 | 1.03 (0.72 – 1.48) |
|  | mRNA-1273 | <11 | 1.24 (0.44 – 3.46) | 48 | 1.01 (0.68 – 1.51) |
| Myocarditis/Pericarditis: All Care Settings (18-45 years of age) | BNT162b2 | <11 | 2.65 (0.95 – 7.41) | 38 | 1.96 (1.32 – 2.93) |
|  | mRNA-1273 | <11 | **7.22\* (2.58 – 20.19)** | 21 | 2.09 (1.16 – 3.77) |
| Myocarditis/Pericarditis: All Care Settings (46-64 years of age) | BNT162b2 | <11 | 1.22 (0.45 – 3.28) | 35 | 1.23 (0.89 – 1.71) |
|  | mRNA-1273 | <11 | **6.61\* (3.26 – 13.41)** | 27 | 1.45 (1.00 – 2.10) |
| Myocarditis/Pericarditis: IP/OP-ED (18-45 years of age) | BNT162b2 | 0 | NA | 15 | 2.5 (1.28 – 4.85) |
|  | mRNA-1273 | 0 | NA | <11 | **3.63\* (1.62 – 8.15)** |
| Myocarditis/Pericarditis: IP/OP-ED (46-64 years of age) | BNT162b2 | <11 | 4.66 (0.84 – 25.98) | <11 | 1.02 (0.51 – 2.04) |
|  | mRNA-1273 | <11 | **11.35\* (4.06 – 31.74)** | <11 | 2.03 (1.09 – 3.77) |
| Narcolepsy | BNT162b2 | 12 | 1.12 (0.39 – 3.23) | 84 | 0.8 (0.63 – 1.02) |
|  | mRNA-1273 | <11 | 1.05 (0.47 – 2.34) | 52 | 0.88 (0.59 – 1.31) |
| Non-hemorrhagic Stroke | BNT162b2 | <11 | 0.53 (0.24 – 1.18) | 92 | 0.82 (0.55 – 1.24) |
|  | mRNA-1273 | <11 | 1.69 (0.91 – 3.13) | 56 | 0.77 (0.59 – 0.99) |
| Pulmonary Embolism | BNT162b2 | 28 | 1.08 (0.51 – 2.28) | 281 | 1 (0.89 – 1.13) |
|  | mRNA-1273 | 14 | 1.06 (0.64 – 1.77) | 167 | 0.97 (0.78 – 1.20) |
| Transverse Myelitis | BNT162b2 | 0 | NA | <11 | 1.06 (0.32 – 3.59) |
|  | mRNA-1273 | 0 | NA | <11 | 1.32 (0.39 – 4.44) |
| Unusual Site Thrombosis with Thrombocytopenia Syndrome | BNT162b2 | <11 | **8.43\* (1.51 – 46.96)** | <11 | 0.73 (0.22 – 2.47) |
|  | mRNA-1273 | 0 | NA | <11 | 1.01 (0.30 – 3.39) |

*\*Indicates the adverse event for the specific vaccine brand has signaled in the analyses.*
*Cells with ‘NA’ indicate adverse events for which an IRR (95% CI) could not be estimated due to zero observed cases.*
*Cell sizes 1-10 were masked for confidentiality.*

**Supplemental Table 5.** Incident Rate Ratio (IRR) and 95% Confidence Interval (CI) of Adverse Events Following a Third Dose of COVID-19 mRNA Vaccine Among Adults 65 Years and Older Enrolled in Medicare, Stratified by Prior COVID-19 Diagnosis.

| Adverse Events | Vaccine Brand | With Prior COVID-19 Diagnosis |  |  | Without Prior COVID-19 Diagnosis |  |  |
| --- | --- | --- | --- | --- | --- | --- | --- |
|  |  | No. Observed Cases | Observed Person-Time (Years) | IRR (95% CI) | No. Observed Cases | Observed Person-Time (Years) | IRR (95% CI) |
| Acute Myocardial Infarction | BNT162b2 | 274 | 14,705 | **1.15\* (1.02 – 1.29)** | 2204 | 199,895 | 0.86 (0.83 – 0.90) |
|  | mRNA-1273 | 216 | 11,396 | **1.33\* (1.16 – 1.52)** | 2,218 | 205,457 | 0.87 (0.84 – 0.91) |
| Anaphylaxis | BNT162b2 | 0 | 1,140 | 0 (0.00 – 41.98) | <11 | 15,198 | 2.26 (0.47 – 6.61) |
|  | mRNA-1273 | 0 | 888 | 0 (0.00 – 49.92) | 0 | 15,647 | 0 (0.00 – 2.66) |
| Appendicitis | BNT162b2 | 21 | 21,878 | 1.31 (0.81 – 2.01) | 218 | 297,048 | 0.93 (0.81 – 1.06) |
|  | mRNA-1273 | <11 | 16,796 | 0.85 (0.42 – 1.52) | 247 | 303,425 | 1.02 (0.89 – 1.15) |
| Bell's Palsy | BNT162b2 | 60 | 22,881 | 1.14 (0.87 – 1.47) | 743 | 311,158 | **1.11\* (1.03 – 1.19)** |
|  | mRNA-1273 | 48 | 17,662 | 1.23 (0.91 – 1.63) | 674 | 318,725 | 0.99 (0.91 – 1.06) |
| Common Site Thrombosis with Thrombocytopenia Syndrome | BNT162b2 | 83 | 15,514 | 1.23 (0.98 – 1.52) | 654 | 208,777 | 0.88 (0.81 – 0.95) |
|  | mRNA-1273 | 57 | 12,076 | 1.19 (0.90 – 1.54) | 651 | 215,843 | 0.86 (0.80 – 0.93) |
| Deep Vein Thrombosis | BNT162b2 | 261 | 14,696 | 1 (0.88 – 1.13) | 2,037 | 200,912 | 0.76 (0.73 – 0.79) |
|  | mRNA-1273 | 166 | 11,516 | 0.94 (0.80 – 1.09) | 1,860 | 207,301 | 0.7 (0.66 – 0.73) |
| Disseminated Intravascular Coagulation | BNT162b2 | 0 | 14,267 | 0 (0.00 – 0.54) | 63 | 194,039 | 0.91 (0.70 – 1.16) |
|  | mRNA-1273 | <11 | 10,888 | 1.53 (0.61 – 3.15) | 46 | 197,137 | 0.67 (0.49 – 0.89) |
| Encephalitis or Encephalomyelitis | BNT162b2 | <11 | 21,208 | 1.33 (0.27 – 3.88) | 35 | 295,147 | 1.38 (0.96 – 1.92) |
|  | mRNA-1273 | <11 | 15,949 | 1.3 (0.16 – 4.68) | 31 | 296,728 | 1.23 (0.83 – 1.74) |
| Guillain-Barré Syndrome | BNT162b2 | 0 | 21,093 | 0 (0.00 – 4.22) | 12 | 290,803 | 0.92 (0.48 – 1.61) |
|  | mRNA-1273 | 0 | 15,939 | 0 (0.00 – 5.36) | <11 | 294,353 | 0.75 (0.36 – 1.39) |
| Hemorrhagic Stroke | BNT162b2 | 40 | 14,759 | 1.06 (0.76 – 1.45) | 353 | 199,223 | 0.85 (0.77 – 0.95) |
|  | mRNA-1273 | 26 | 11,382 | 1 (0.65 – 1.47) | 344 | 204,032 | 0.84 (0.75 – 0.93) |
| Immune Thrombocytopenia | BNT162b2 | 37 | 22,705 | **1.66\* (1.17 – 2.29)** | 296 | 305,480 | 1.07 (0.95 – 1.20) |
|  | mRNA-1273 | 17 | 17,624 | 1.02 (0.59 – 1.63) | 305 | 314,669 | 1.08 (0.96 – 1.20) |
| Myocarditis/Pericarditis | BNT162b2 | 22 | 11,554 | **1.89\* (1.18 – 2.86)** | 139 | 154,087 | 1.02 (0.86 – 1.21) |
|  | mRNA-1273 | 13 | 9,000 | 1.53 (0.81 – 2.62) | 131 | 158,882 | 0.94 (0.79 – 1.12) |
| Narcolepsy | BNT162b2 | 12 | 22,422 | 1.26 (0.65 – 2.19) | 123 | 302,017 | 1.1 (0.92 – 1.32) |
|  | mRNA-1273 | <11 | 17,356 | 0.72 (0.23 – 1.68) | 88 | 310,198 | 0.77 (0.62 – 0.95) |
| Non-hemorrhagic Stroke | BNT162b2 | 158 | 14,825 | 1 (0.85 – 1.17) | 1,269 | 200,076 | 0.74 (0.70 – 0.78) |
|  | mRNA-1273 | 101 | 11,486 | 0.93 (0.76 – 1.13) | 1,288 | 205,633 | 0.76 (0.72 – 0.80) |
| Pulmonary Embolism | BNT162b2 | 182 | 14,951 | **1.36\* (1.17 – 1.58)** | 1,567 | 202,824 | 1.02 (0.97 – 1.07) |
|  | mRNA-1273 | 137 | 11,671 | **1.43\* (1.20 – 1.69)** | 1,512 | 209,413 | 0.98 (0.93 – 1.03) |
| Transverse Myelitis | BNT162b2 | <11 | 21,625 | 2.21 (0.27 – 7.99) | 15 | 295,496 | 1.51 (0.85 – 2.50) |
|  | mRNA-1273 | <11 | 16,502 | 3.19 (0.39 – 11.53) | <11 | 301,087 | 0.4 (0.11 – 1.02) |
| Unusual Site Thrombosis with Thrombocytopenia Syndrome | BNT162b2 | <11 | 15,472 | 2.63 (0.72 – 6.74) | 24 | 208,130 | 1.17 (0.75 – 1.73) |
|  | mRNA-1273 | <11 | 11,997 | 0.83 (0.02 – 4.65) | 19 | 214,869 | 0.88 (0.53 – 1.38) |

*\*Indicates the adverse event for the specific vaccine brand has signaled in the analyses.*
*Medicare database contains data through March 2022.*
*Cell sizes 1-10 were masked for confidentiality.*

**Supplemental Table 6.** Number of COVID-19 Homologous and Heterologous Third Vaccine Doses Administered Following Primary Series.

| Vaccine Brand | Optum | Carelon Research | CVS Health | CMS |
| --- | --- | --- | --- | --- |
|  | No. Vaccinees (%) | No. Vaccinees (%) | No. Vaccinees (%) | No. Vaccinees (%) |
| **Primary Series: BNT162b2** | **1,182,306** | **1,326,121** | **1,023,199** | **5,208,339** |
| Total third vaccine doses among BNT162b2 primary series recipients | 1,182,306 (100.00) | 1,326,121 (100.00) | 1,023,199 (100.00) | 3,243,792 (100.00) |
| Ad26.COV2.S booster doses | -- | -- | -- | 3,301 (0.10) |
| BNT162b2 third vaccine doses | 1,062,247 (89.85) | 1,198,742 (90.39) | 903,546 (88.31) | 3,136,719 (96.70) |
| mRNA-1273 third vaccine doses | 120,059 (10.15) | 127,379 (9.61) | 119,653 (11.69) | 103,772 (3.20) |
| **Primary Series: mRNA-1273** | **700,210** | **768,750** | **585,837** | **5,233,843** |
| Total third vaccine doses among mRNA-1273 primary series recipients | 700,210 (100.00) | 768,750 (100.00) | 585,837 (100.00) | 3,370,141 (100.00) |
| Ad26.COV2.S booster doses | -- | -- | -- | 6,259 (0.19) |
| BNT162b2 third vaccine doses | 72,499 (10.35) | 88,046 (11.45) | 62,976 (10.75) | 149,253 (4.43) |
| mRNA-1273 third vaccine doses | 627,711 (89.65) | 680,704 (88.55) | 522,861 (89.25) | 3,214,629 (95.39) |
| **Primary Series: Ad26.COV2.S** | **37,610** | **42,855** | **26,700** | **541,723** |
| Total third vaccine doses among Ad26.COV2.S primary series recipients | 37,610 (100.00) | 42,855 (100.00) | 26,700 (100.00) | 211,684 (100.00) |
| Ad26.COV2.S booster doses | 25,480 (67.75) | 35,565 (82.99) | 21,411 (80.19) | 80,147 (37.86) |
| BNT162b2 third vaccine doses | 6,318 (16.80) | 3,715 (8.67) | 2,340 (8.76) | 50,890 (24.04) |
| mRNA-1273 third vaccine doses | 5,812 (15.45) | 3,575 (8.34) | 2,949 (11.04) | 80,647 (38.10) |

## Ethics Statement

As a public health surveillance activity, this study was not subject to requirements for Institutional Review Board review.

## Authorship Contributions

All authors contributed to the concept and design of the study and interpretation of the data. A. Shoaibi drafted the manuscript and all authors reviewed it. K. Matuska, H. Lyu, R. McEvoy, Z. Wan, M. Hu, D. Beachler, A. Secora, N. Selvam, D. Djibo, C. McMahill-Walraven, J. Seeger, K. Amend, J. Song, R. Clifford, R. Forshee, and S. Anderson acquired and managed the data and performed analyses. The views expressed here are those of the authors and not necessarily those of the U.S. Food and Drug Administration, the Centers for Medicare and Medicaid Services, the Department of Health and Human Services, CVS Health/Aetna, Carelon Research, IQVIA, or Optum Epidemiology. This work has not been presented publicly, previously published, or is under current consideration by another journal.

## Data Availability

The study protocol was publicly posted as referenced in the manuscript before data analyses, and related documents can be made available where needed by contacting the corresponding author. De-identified participant data will not be shared without approval from the data partners.

## Acknowledgments

We would like to thank Gyanada Acharya, Derick Ambarsoomzadeh, Mahasweta Mitra, Michelle Ondari, Ellie Smith, and Zoe Wu from Acumen LLC; Shiva Vojjala, Ramya Avula, Shiva Chaudhary, Qian Si, Dianna Hayden, Shanthi P Sagare, Ramin Riahi, Brian Greenwald, and Grace Stockbower from Carelon Research; Michael Goodman, Michael Bruhn, and Ruth Weed from IQVIA; Anne Marie Kline, Nancy B. Shaik, Ana M. Martinez-Baquero, Smita Bhatia, Carla Brannan, Vaibhav Sharma, Eugenio Abente, and Jonathan Deshazo from CVS Health; and Grace Yang, Sarah Sargen, Alexandra Stone, Wafa Tarazi, Megan Ketchell, Kathryn Federici, Amaka Ume, Emily Myers, Eli Wolter, Jackson Slaney, Bobby Smith, Lauren Peetluk, and Elizabeth Bell from Optum for their assistance with data validation, analysis, manuscript preparation, and project coordination.

